# Examining the Pervasiveness of the Impact of COVID-19 and Associated Lockdown Policies on Pneumococcal Conjugate Vaccine (PCV) Coverage

**DOI:** 10.1101/2025.06.23.25330136

**Authors:** Ruoxuan Wang, Bryan N. Patenaude

## Abstract

The COVID-19 pandemic resulted in significant reductions in the immunization rate of routine pneumococcal conjugate vaccines (PCV) worldwide due to disruptions in vaccine supply chains, shortages of healthcare workers, facility closure or overcrowding, and parents’ concerns that their children would become infected with COVID-19 (SeyedAlinaghi et al., 2022; Basu et al., 2023; Lan et al., 2023; Hora et al., 2023). Our research utilizes a longitudinal difference-in-difference approach, to quantify the immediate impact that the COVID-19 pandemic and country-specific pandemic response measures had on routine PCV vaccine coverage, and the relative speed at which routine PCV coverage rebounded toward pre-pandemic levels in the post-2020 period between countries that adopted specific types of lockdown measures and those that did not. We also examine whether immunization catch-up campaign capacity and preparedness mitigate the impact of COVID-19 and pandemic response policies or resulted in faster recovery of PCV vaccination to pre-pandemic levels. We utilize data from the World Health Organization (2023) on PCV coverage from 2014 to 2023 and Hale et al. (2021) on country-specific lockdown policy enactment during the COVID-19 pandemic for 106 countries. Across all countries, the pandemic had a detrimental impact on PCV coverage. Pandemic lockdown policies, however, had a mixed impact on PCV coverage. Countries that enacted school closures and strict stay-at-home orders saw no significant differences in the effect of COVID-19 on PCV coverage compared with countries that did enact school closures, and these countries did not experience any differences in the speed of PCV vaccination coverage recovery post-pandemic than countries that did not enact school closures. Likewise, countries that enacted workplace closures showed no differences in the immediate effect of COVID-19 on PCV coverage compared with countries that did not enact workplace closures. However, countries that enacted workplace closures recovered PCV coverage in the post-period slightly faster than countries that did not enact workplace closures (0.893 percentage points per year, 95% CI = [0.078, 1.709]), even after controlling for the severity of the pandemic, indicating that workplace closure had the lowest longer-term disruptive impact on PCV coverage among all the lockdown policies employed. While catch-up campaigns resulted in significant coverage gains in the pre-pandemic period (1.50 percentage points per year 95% CI= [0.202, 2.979]), experience with implementing successful catchup campaigns did not mitigate the impact of COVID-19 or lockdown policy on PCV coverage. These findings suggest that capacity and training alone is insufficient for health systems preparedness, and pandemic response plans need to ensure adequate healthcare workforce, supply chain security, and health financing for impact mitigation in order to protect routine vaccination rates from decreasing during future pandemics and other public health disasters.

## Introduction

Prior research has shown that the COVID-19 pandemic disrupted the timely delivery of routine immunizations, reducing coverage levels and increasing the risk of outbreaks of vaccine-preventable diseases such as pneumococcal disease (Basu et al., 2023; Bertran et al, 2022; Lan et al., 2023). Basu et al. (2023) conducted a global evaluation of the impact of the COVID-19 pandemic on supply chain disruptions for routine immunizations, finding that the COVID-19 pandemic and associated response resulted in a 9% decrease in vaccine supply for polio and human papillomavirus vaccines (HPV) and a 2-3% reduction in Measles-containing-vaccine second-dose (MCV2) coverage. Additionally, the global coverage of diphtheria, tetanus, and pertussis (DTP) vaccine decreased from 86% to 83% between 2019 and 2020, as a result of the COVID-19 pandemic (The Lancet Child Adolescent Health, 2021). For pneumococcal disease, Bertran et al. (2022) showed that PCV immunization disruptions particularly impacted populations already vulnerable to vaccine-preventable illness and found that from July to December 2021, the incidence of pneumococcal disease among children below age 15 increased from the pre-pandemic level of 1.43/100,000 children to a level of 1.96 per 100,000 children in England (Bertran et al., 2022). At a global scale, Ngigi et al. (2024) show that the COVID-19 pandemic resulted in a 21.6% decrease in PCV vaccine coverage overall and a 20.1% decline in the receipt of a second dose, highlighting the statistically significant and negative impact of the COVID-19 pandemic on PCV vaccination rates, globally.

In addition to vaccine supply chain disruptions, several other factors contributed to a decline in routine immunization coverage during the COVID-19 pandemic. On the supply side, shortages of nurses and primary healthcare providers delayed routine vaccination administration because these workers were re-directed to COVID-19 outbreak response activities (Hora et al., 2023). Moreover, lockdowns and stay-at-home requirements resulted in delays in scheduling timely vaccination clinic visits in accordance with country vaccine administration schedules (SeyedAlinaghi et al., 2022). On the demand side, many parents avoided or delayed taking their children to routine vaccination appointments during the COVID-19 pandemic due to concerns that their children would become exposed to the SARS-CoV-2 virus (SeyedAlinaghi et al., 2022). Basu et al. (2023) point out that declines in pediatric vaccine coverage during the COVID-19 pandemic contributed to a substantial increase in the number of zero-dose children worldwide, defined by Gavi as children who have never received a single dose of DTP vaccine (Basu et al., 2023). In sub-Saharan Africa, the number of zero dose children rose from 7.1 million to 7.7 million, and in Southeast Asia rose from 2.0 to 4.1 million children between 2019 and 2022 (Basu et al., 2023).

Despite well documented trends in the reduction of routine pediatric immunization coverage during the COVID-19 pandemic, there is limited evidence on heterogeneity in the pervasiveness of the impact or whether specific infection mitigation actions taken by countries resulted in more or less pervasive coverage declines in routine immunization coverage. To fill this gap, our study quantifies the pervasiveness of the impact that the COVID-19 pandemic as well as pandemic infection control policies (school closing, workplace closing, and stay-at-home requirements) had on PCV vaccine coverage. We also examine whether or not implementing a recent pre-pandemic immunization catch-up campaign may have mitigated the pervasiveness of adverse impacts, due to delivery system preparedness to respond to reductions in coverage. Our findings highlight the importance of health systems preparedness in mitigating and resolving potential negative externalities on routine vaccination coverage due to public health emergencies, such as the COVID-19 pandemic.

## Methods

Our analysis utilizes PCV coverage reported by the World Health Organization from 2014 to 2023 (World Health Organization, 2023) as the primary outcome of the analysis. Data on country-specific lockdown policies were obtained from Hale et al. (2021) and coded as binary indicators at the country-level for having enacted the policy between March of 2020 and December of 2021. We then employed a difference-in-difference analysis with both pre- and post-period trends, to examine the impact and pervasiveness of the impact that residing in a country that enacted school closures, workplace closures, or stay-at-home requirements, had on PCV vaccine coverage compared to countries that did not enact such policies. We employ a random effects panel data difference-in-difference model in order to assess the mitigating effect of time-invariant characteristics, including whether or not the country had recent pre-pandemic experience in implementing catch-up vaccination campaigns, on the level and pervasiveness of the impact on coverage. Using data from the World Bank (2025) and the World Health Organization (2025), we incorporate gross domestic product per capita and number of COVID-19 cases as covariates into the regression models to control for the strength of the economy as well as the severity of the pandemic.

Specifically, Models 1, 2, and 3 examine the association between pandemic lockdown policies and PCV coverage by comparing national vaccination rate between countries with and without these policies. Model 4 analyzes whether post-pandemic PCV coverage differs between countries with and without pre-pandemic vaccine catch-up campaigns. Models 5, 6, and 7 compare whether countries that implemented both pre-pandemic vaccine catch-up campaigns and COVID-19 lockdown policies experienced a faster post-pandemic recovery in their PCV coverage than countries that implemented either or neither of these interventions. The (a) versions of the model do not contain controls for per capita GDP and COVID-19 cases while the (b) versions add in these controls.

### Regression Equations

1. **Model 1(a):** PCV_coverage_it = β + β (YEAR_it) + β (PRE_YEAR_C1M_it) + β (C1M_it) + β (POST_YEAR_it) + β (POST_YEAR_C1M_it) + ε_it
2. **Model 1(b):** PCV_coverage_it = β + β (YEAR_it) + β (PRE_YEAR_C1M_it) + β (C1M_it) + β (POST_YEAR_it) + β (POST_YEAR_C1M_it) + β (GDPPC_it) + β (COVID19_Cases_it) + ε_it
3. **Model 2(a):** PCV_coverage_it = β + β (YEAR_it) + β (PRE_YEAR_C6M_it) + β (C6M_it) + β (POST_YEAR_it) + β (POST_YEAR_C6M_it) + ε_it
4. **Model 2(b):** PCV_coverage_it = β + β (YEAR_it) + β (PRE_YEAR_C6M_it) + β (C6M_it) + β (POST_YEAR_it) + β (POST_YEAR_C6M_it) + β (GDPPC_it) + β (COVID19_Cases_it) + ε_it
5. **Model 3(a):** PCV_coverage_it = β + β (YEAR_it) + β (PRE_YEAR_C2M_it) + β (C2M_it) + β (POST_YEAR_it) + β (POST_YEAR_C2M_it) + ε_it
6. **Model 3(b):** PCV_coverage_it = β + β (YEAR_it) + β (PRE_YEAR_C2M_it) + β (C2M_it) + β (POST_YEAR_it) + β (POST_YEAR_C2M_it) + β (GDPPC_it) + β (COVID19_Cases_it) + ε_it
7. **Model 4(a):** PCV_coverage_it = β + β (YEAR_it) + β (PRE_YEAR_CATCHUP_it) + β (CATCHUP_it) + β (POST_YEAR_it) + β (POST_YEAR_CATCHUP_it) + ε_it
8. **Model 4(b):** PCV_coverage_it = β + β (YEAR_it) + β (PRE_YEAR_CATCHUP_it) + β (CATCHUP_it) + β (POST_YEAR_it) + β (POST_YEAR_CATCHUP_it) + β (GDPPC_it) + β (COVID19_Cases_it) + ε_it
9. **Model 5(a):** PCV_coverage_it = β + β (YEAR_it) + β (PRE_YEAR_C1M_it) + β (PRE_YEAR_CATCHUP_it) + β (PRE_YEAR_C1M_CATCHUP_it) + β (C1M_it) + β (CATCHUP_it) + β (POST_YEAR_it) + β (POST_YEAR_C1M_it) + β (POST_YEAR_CATCHUP_it) + β (POST_YEAR_C1M_CATCHUP_it) + ε_it
10. **Model 5(b):** PCV_coverage_it = β + β (YEAR_it) + β (PRE_YEAR_C1M_it) + β (PRE_YEAR_CATCHUP_it) + β (PRE_YEAR_C1M_CATCHUP_it) + β (C1M_it) + β (CATCHUP_it) + β (POST_YEAR_it) + β (POST_YEAR_C1M_it) + β (POST_YEAR_CATCHUP_it) + β (POST_YEAR_C1M_CATCHUP_it) + β (GDPPC_it) + β (COVID19_Cases_it) + ε_it
11. **Model 6(a):** PCV_coverage_it = β + β (YEAR_it) + β (PRE_YEAR_C2M_it) + β (PRE_YEAR_CATCHUP_it) + β (PRE_YEAR_C2M_CATCHUP_it) + β (C2M_it) + β (CATCHUP_it) + β (POST_YEAR_it) + β (POST_YEAR_C2M_it) + β (POST_YEAR_CATCHUP_it) + β (POST_YEAR_C2M_CATCHUP_it) + ε_it
12. **Model 6(b):** PCV_coverage_it = β + β (YEAR_it) + β (PRE_YEAR_C2M_it) + β (PRE_YEAR_CATCHUP_it) + β (PRE_YEAR_C2M_CATCHUP_it) + β (C2M_it) + β (CATCHUP_it) + β (POST_YEAR_it) + β (POST_YEAR_C2M_it) + β (POST_YEAR_CATCHUP_it) + β (POST_YEAR_C2M_CATCHUP_it) + β (GDPPC_it) + β (COVID19_Cases_it) + ε_it
13. **Model 7(a):** PCV_coverage_it = β + β (YEAR_it) + β (PRE_YEAR_C6M_it) + β (PRE_YEAR_CATCHUP_it) + β (PRE_YEAR_C6M_CATCHUP_it) + β (C6M_it) + β (CATCHUP_it) + β (POST_YEAR_it) + β (POST_YEAR_C6M_it) + β (POST_YEAR_CATCHUP_it) + β (POST_YEAR_C6M_CATCHUP_it) + ε_it
14. **Model 7(b):** PCV_coverage_it = β + β (YEAR_it) + β (PRE_YEAR_C6M_it) + β (PRE_YEAR_CATCHUP_it) + β (PRE_YEAR_C6M_CATCHUP_it) + β (C6M_it) + β (CATCHUP_it) + β (POST_YEAR_it) + β (POST_YEAR_C6M_it) + β (POST_YEAR_CATCHUP_it) + β (POST_YEAR_C6M_CATCHUP_it) + β (GDPPC_it) + β (COVID19_Cases_it) + ε_it

**Table 1:**
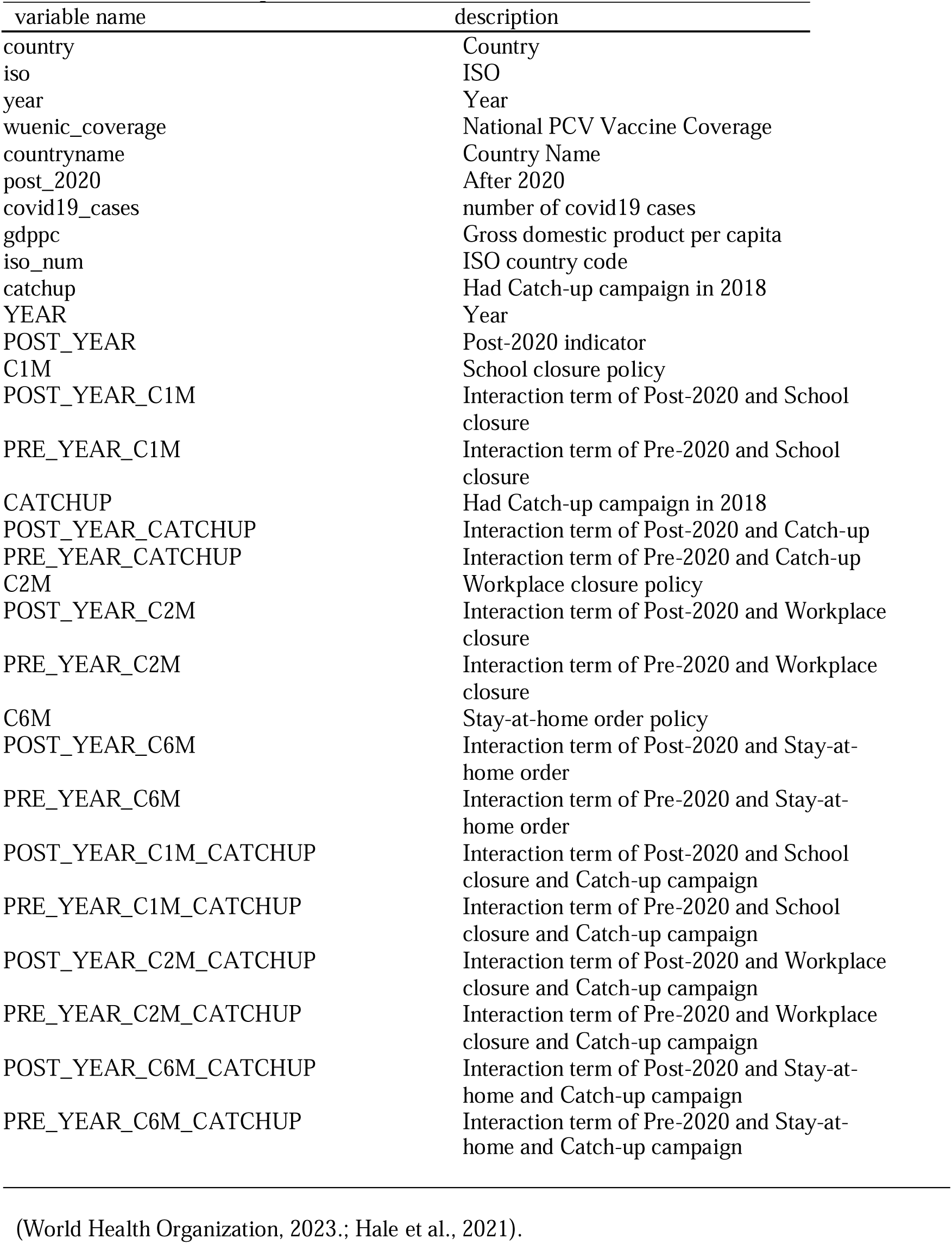
Variable Descriptions.

## Results

The COVID-19 pandemic had an adverse impact on PCV coverage for all countries and resulted in an annual decrease of coverage by 0.69 to 1.62 percentage points. Our analysis shows that pandemic lockdown policies had varying effects on post-pandemic PCV coverage. For example, countries that enacted strict stay-at-home orders experienced no differences in the effect of COVID-19 on PCV coverage compared with countries that did not enact stay-at-home orders, which were Benin, Botswana, Burundi, Cameroon, Central African Republic, Costa Rica, Ethiopia, Japan, Iceland, Kiribati, Malawi, Mozambique, Nicaragua, Norway, Oman, Sierra Leone, Singapore, and Zambia (Hale et al., 2021) and also exhibited no differences in the speed of recovery post-COVID compared with countries that did not enact stay-at-home orders.

**Figure 1:**
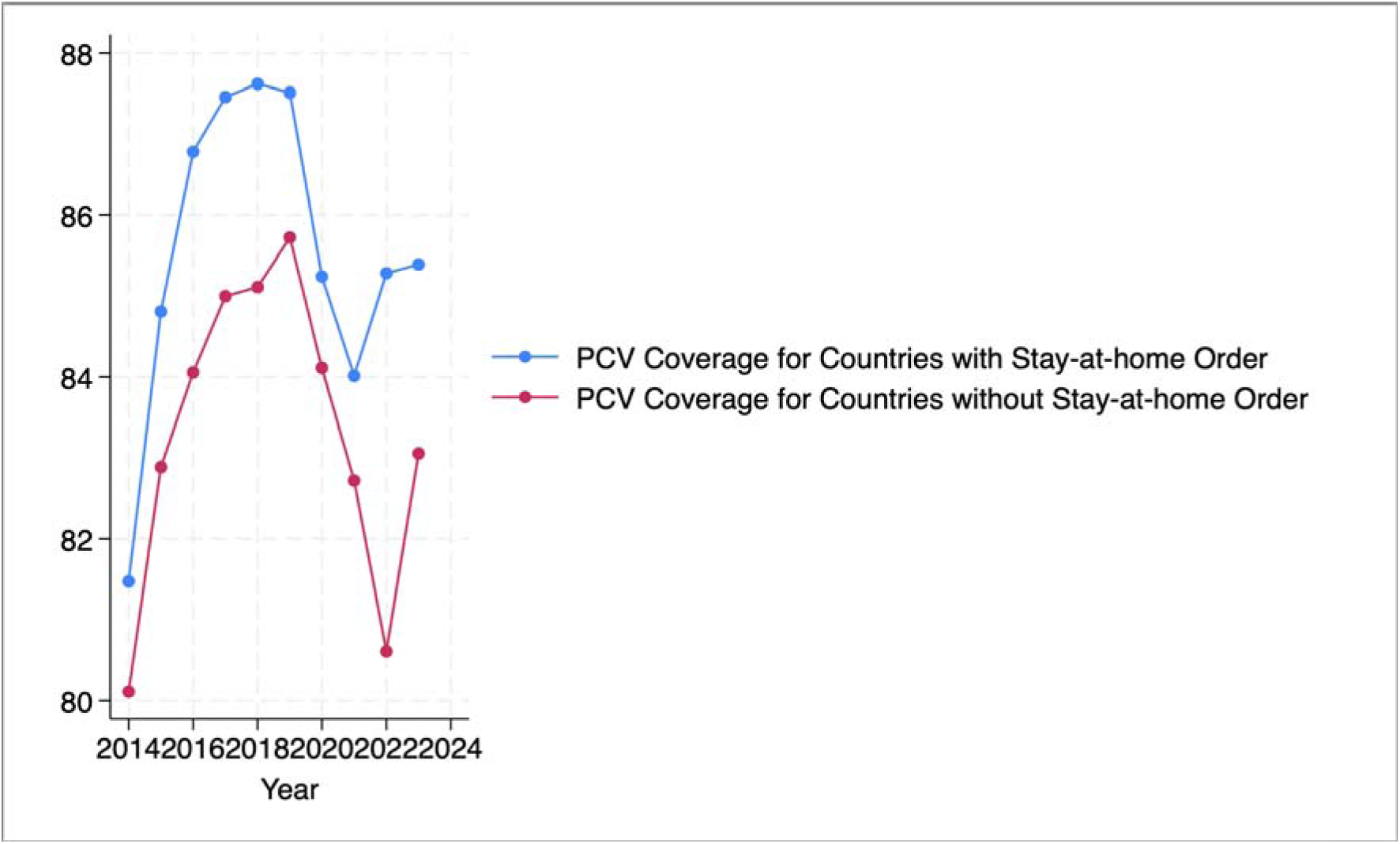
PCV Coverage in Countries with and without Stay-at-home Requirements:

Similarly, countries that enacted workplace closures saw no differences in the effect of COVID-19 on PCV coverage compared with countries that did not enact workplace closures, which were Botswana, Kiribati, Burundi, Malawi, Mozambique, Nicaragua, Niger, Senegal, Yemen, and Zambia. However, there is some modest evidence that countries that enacted workplace closures recovered PCV coverage in the post-period faster than countries that did not enact workplace closures, with countries enacting workplace closures recovering PCV coverage at a rate of 0.893 percentage points per year faster than countries that did not enact workplace closures. This finding also demonstrates that workplace closure policy had the lowest longer-term impacts on PCV coverage among all the lockdown strategies employed.

**Figure 2:**
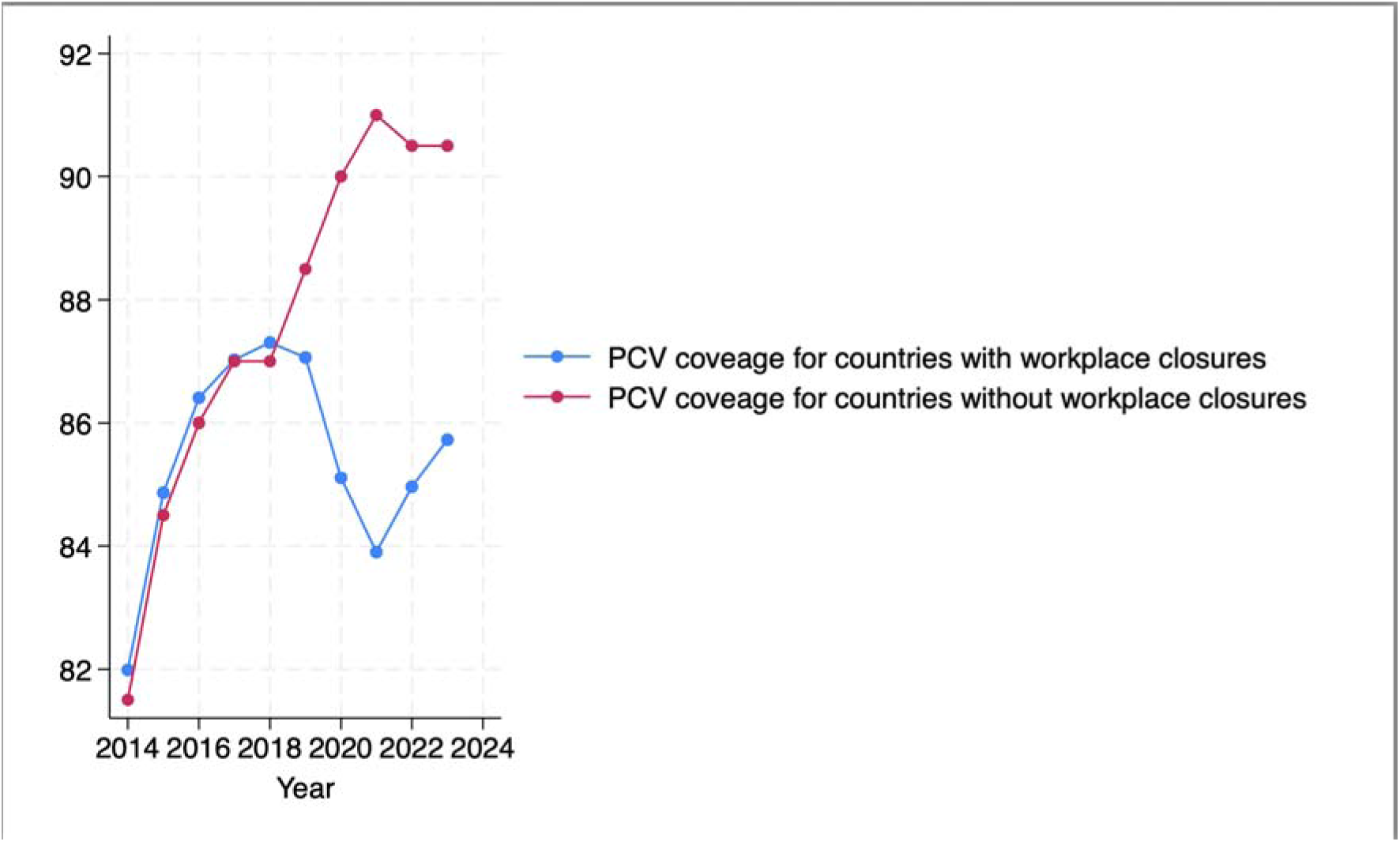
PCV Coverage in Countries with and without Workplace Closures:

Furthermore, countries that enacted school closures experienced no differences in the effect of COVID-19 on PCV coverage compared with countries that did enact school closures and saw no differences in the speed of recovery post-covid than countries that did not enact school closures. Because only two countries—Singapore and Burundi—did not enact school closure during the pandemic and both of these countries implemented vaccine catch-up campaigns in 2018, the parallel trends assumption for the difference-in-difference is violated in the period immediately leading up to the enactment of school closures (Barikumutima, 2019). Instead of being able to speak to school closures in particular, we instead classified countries based on whether they implemented pre-pandemic vaccine campaigns to examine whether experience in administering catch-up campaigns mitigated the impact that the COVID-19 pandemic had on PCV coverage globally, by building capacity to and experience to roll out catch-up vaccination efforts.

**Figure 3:**
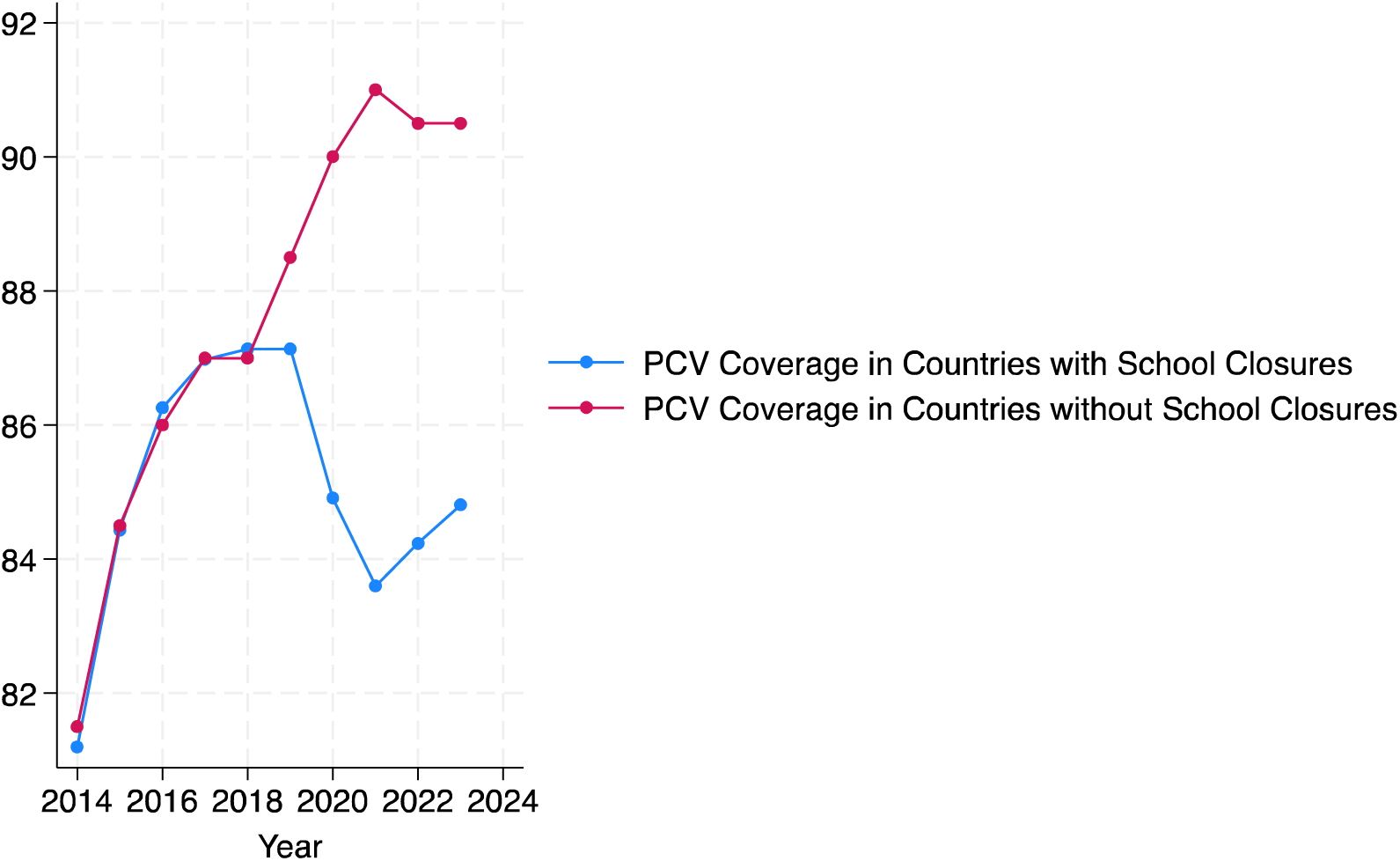
PCV Coverage in Countries with and without School Closures:

Although countries that enacted catchup campaigns in the pre-COVID period had lower coverage on average, they saw faster increases in PCV coverage in the pre-period of 1.5 percentage points per year than countries that did not have campaigns. Nonetheless, experience with implementing successful catchup campaigns in the pre-period did not result in the mitigation of the impact of COVID-19 or lockdown policies on PCV coverage. No evidence demonstrated that countries that had enacted these campaigns either were less impacted by COVID-19 or lockdown policies or recovered PCV coverage faster from COVID-19 or lockdown policy. Only some initial weak evidence suggested that countries that employed stay at home orders and had catchup campaigns might have seen larger coverage declines in the post-period, but this effect was removed once controlling for the severity of the pandemic and wealth of the country indicating that the initial effect was likely due to the fact that these specific countries were poorer or more severely impacted by COVID-19 than other countries.

**Figure 4:**
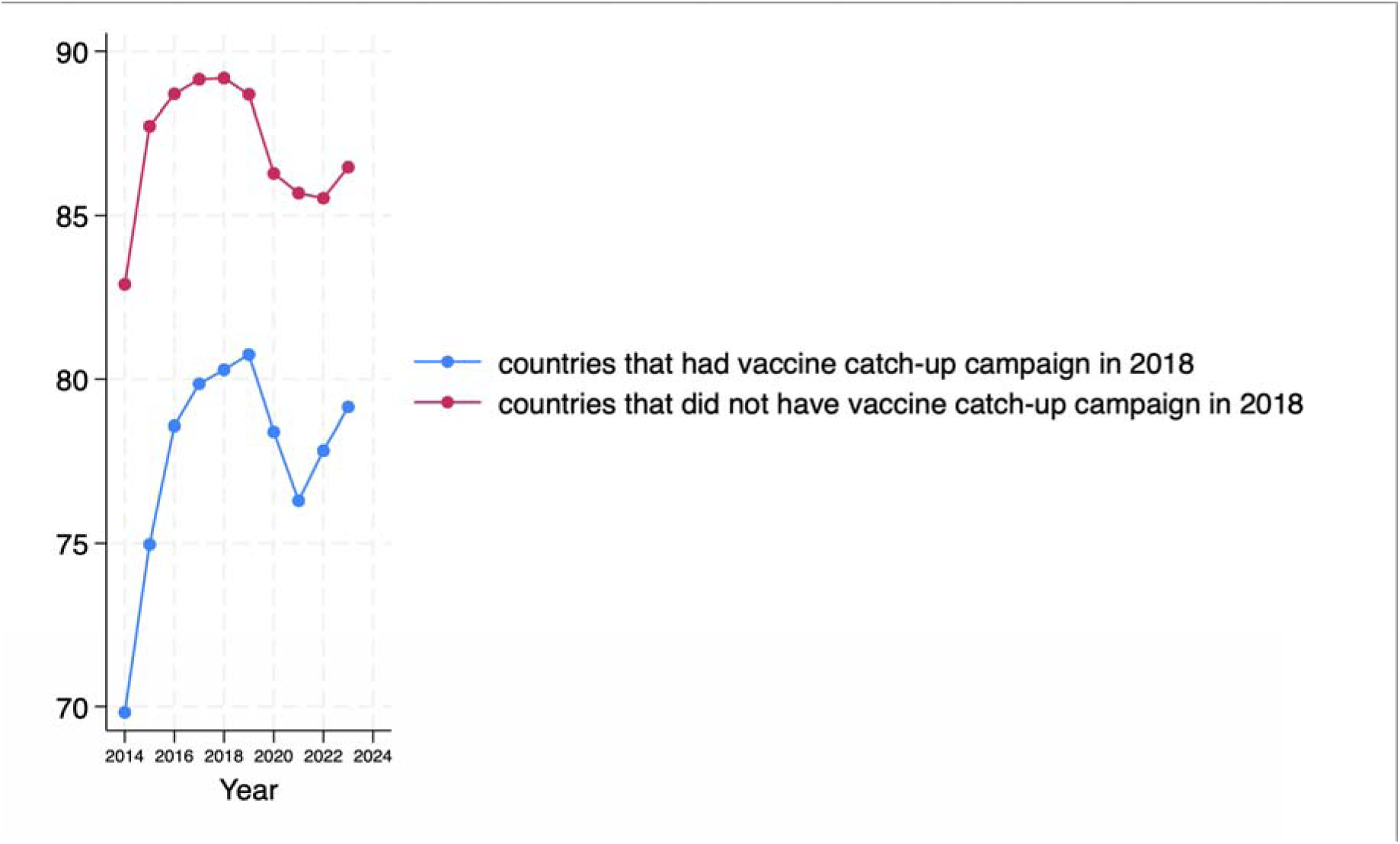
PCV Coverage in Countries with and without Pre-Pandemic Catch-up Campaigns

**Table 2:** Countries with vaccine catchup campaigns in 2018:

| Country | Freq. | Percent | Cum. |
| --- | --- | --- | --- |
| Afghanistan | 10 | 3.57 | 3.57 |
| Benin | 10 | 3.57 | 7.14 |
| Burkina Faso | 10 | 3.57 | 10.71 |
| Burundi | 10 | 3.57 | 14.29 |
| Cameroon | 10 | 3.57 | 17.86 |
| Central African Republic | 10 | 3.57 | 21.43 |
| Djibouti | 10 | 3.57 | 25.00 |
| Ethiopia | 10 | 3.57 | 28.57 |
| Gambia | 10 | 3.57 | 32.14 |
| Ghana | 10 | 3.57 | 35.71 |
| Kenya | 10 | 3.57 | 39.29 |
| Liberia | 10 | 3.57 | 42.86 |
| Madagascar | 10 | 3.57 | 46.43 |
| Malawi | 10 | 3.57 | 50.00 |
| Mali | 10 | 3.57 | 53.57 |
| Mauritania | 10 | 3.57 | 57.14 |
| Mozambique | 10 | 3.57 | 60.71 |
| Niger | 10 | 3.57 | 64.29 |
| Pakistan | 10 | 3.57 | 67.86 |
| Rwanda | 10 | 3.57 | 71.43 |
| Senegal | 10 | 3.57 | 75.00 |
| Sierra Leone | 10 | 3.57 | 78.57 |
| Singapore | 10 | 3.57 | 82.14 |
| Togo | 10 | 3.57 | 85.71 |
| Uganda | 10 | 3.57 | 89.29 |
| Yemen | 10 | 3.57 | 92.86 |
| Zambia | 10 | 3.57 | 96.43 |
| Zimbabwe | 10 | 3.57 | 100.00 |
| Total | 280 | 100.00 |  |

**Table 3:** Countries without pre-pandemic vaccine catch-up campaigns in 2018:

| Country | Freq. | Percent | Cum. |
| --- | --- | --- | --- |
| Albania | 10 | 1.28 | 1.28 |
| Andorra | 10 | 1.28 | 2.56 |
| Angola | 10 | 1.28 | 3.85 |
| Argentina | 10 | 1.28 | 5.13 |
| Australia | 10 | 1.28 | 6.41 |
| Azerbaijan | 10 | 1.28 | 7.69 |
| Bahamas | 10 | 1.28 | 8.97 |
| Bahrain | 10 | 1.28 | 10.26 |
| Barbados | 10 | 1.28 | 11.54 |
| Belgium | 10 | 1.28 | 12.82 |
| Bolivia (Plurinational State of) | 10 | 1.28 | 14.10 |
| Botswana | 10 | 1.28 | 15.38 |
| Brazil | 10 | 1.28 | 16.67 |
| Bulgaria | 10 | 1.28 | 17.95 |
| Canada | 10 | 1.28 | 19.23 |
| Chile | 10 | 1.28 | 20.51 |
| Colombia | 10 | 1.28 | 21.79 |
| Congo | 10 | 1.28 | 23.08 |
| Congo, Democratic Republic of the | 10 | 1.28 | 24.36 |
| Costa Rica | 10 | 1.28 | 25.64 |
| Côte d'Ivoire | 10 | 1.28 | 26.92 |
| Denmark | 10 | 1.28 | 28.21 |
| Dominican Republic | 10 | 1.28 | 29.49 |
| Ecuador | 10 | 1.28 | 30.77 |
| El Salvador | 10 | 1.28 | 32.05 |
| Eswatini | 10 | 1.28 | 33.33 |
| Fiji | 10 | 1.28 | 34.62 |
| Finland | 10 | 1.28 | 35.90 |
| France | 10 | 1.28 | 37.18 |
| Germany | 10 | 1.28 | 38.46 |
| Greece | 10 | 1.28 | 39.74 |
| Guatemala | 10 | 1.28 | 41.03 |
| Guyana | 10 | 1.28 | 42.31 |
| Honduras | 10 | 1.28 | 43.59 |
| Hungary | 10 | 1.28 | 44.87 |
| Iceland | 10 | 1.28 | 46.15 |
| Ireland | 10 | 1.28 | 47.44 |
| Israel | 10 | 1.28 | 48.72 |
| Italy | 10 | 1.28 | 50.00 |
| Japan | 10 | 1.28 | 51.28 |
| Kazakhstan | 10 | 1.28 | 52.56 |
| Kiribati | 10 | 1.28 | 53.85 |
| Kuwait | 10 | 1.28 | 55.13 |
| Lao People's Democratic Republic | 10 | 1.28 | 56.41 |
| Latvia | 10 | 1.28 | 57.69 |
| Libya | 10 | 1.28 | 58.97 |
| Luxembourg | 10 | 1.28 | 60.26 |
| Marshall Islands | 10 | 1.28 | 61.54 |
| Mexico | 10 | 1.28 | 62.82 |
| Micronesia (Federated States of) | 10 | 1.28 | 64.10 |
| Moldova, Republic of | 10 | 1.28 | 65.38 |
| Morocco | 10 | 1.28 | 66.67 |
| Netherlands | 10 | 1.28 | 67.95 |
| New Zealand | 10 | 1.28 | 69.23 |
| Nicaragua | 10 | 1.28 | 70.51 |
| Niue | 10 | 1.28 | 71.79 |
| Norway | 10 | 1.28 | 73.08 |
| Oman | 10 | 1.28 | 74.36 |
| Palau | 10 | 1.28 | 75.64 |
| Panama | 10 | 1.28 | 76.92 |
| Paraguay | 10 | 1.28 | 78.21 |
| Peru | 10 | 1.28 | 79.49 |
| Philippines | 10 | 1.28 | 80.77 |
| Qatar | 10 | 1.28 | 82.05 |
| Sao Tome and Principe | 10 | 1.28 | 83.33 |
| Saudi Arabia | 10 | 1.28 | 84.62 |
| Slovakia | 10 | 1.28 | 85.90 |
| South Africa | 10 | 1.28 | 87.18 |
| Sudan | 10 | 1.28 | 88.46 |
| Sweden | 10 | 1.28 | 89.74 |
| Switzerland | 10 | 1.28 | 91.03 |
| Tanzania, United Republic of | 10 | 1.28 | 92.31 |
| Trinidad and Tobago | 10 | 1.28 | 93.59 |
| Turkey | 10 | 1.28 | 94.87 |
| United Arab Emirates | 10 | 1.28 | 96.15 |
| United Kingdom of Great Britain and Northern Ireland | 10 | 1.28 | 97.44 |
| United States of America | 10 | 1.28 | 98.72 |
| Uruguay | 10 | 1.28 | 100.00 |
| Total | 780 | 100.00 |  |

**Table 4:** Nested Regression Results:

|  | (1a) | (1b) | (2a) | (2b) | (3a) | (3b) | (4a) | (4b) | (5a) | (5b) | (6a) | (6b) | (7a) | (7b) |
| --- | --- | --- | --- | --- | --- | --- | --- | --- | --- | --- | --- | --- | --- | --- |
|  | wuenic_coverage | wuenic_coverage | wuenic_coverage | wuenic_coverage | wuenic_coverage | wuenic_coverage | wuenic_coverage | wuenic_coverage | wuenic_coverage | wuenic_coverage | wuenic_coverage | wuenic_coverage | wuenic_coverage | wuenic_coverage |
| YEAR | 2.112**<br>(.928) | .595<br>(1.271) | 1.509***<br>(.541) | .811<br>(.5) | 2.163***<br>(.819) | 2.02*<br>(1.163) | .949***<br>(.306) | .717**<br>(.293) | 2.057**<br>(1.002) | -.034<br>(1.644) | 1.997**<br>(.872) | .974<br>(1.313) | 1.691**<br>(.692) | .78<br>(.764) |
| PRE_YEAR_C1M | -1.044<br>(.972) | .615<br>(1.307) |  |  |  |  |  |  | -1.349<br>(1.032) | .793<br>(1.651) |  |  |  |  |
| C1M | 7.237<br>(6.307) | -.052<br>(10.199) |  |  |  |  |  |  | 9.699<br>(6.538) | -1.647<br>(10.707) |  |  |  |  |
| POST_YEAR | -1.38***<br>(.512) | -.706<br>(.886) | -1.081***<br>(.294) | -.76**<br>(.296) | -1.553***<br>(.396) | -1.619***<br>(.533) | -.767***<br>(.156) | -.686***<br>(.152) | -1.461***<br>(.55) | -.868<br>(1.264) | -1.458***<br>(.454) | -1.224*<br>(.708) | -1.211***<br>(.368) | -.836*<br>(.433) |
| POST_YEAR_C1M | .611<br>(.528) | -.135<br>(.896) |  |  |  |  |  |  | .846<br>(.563) | .197<br>(1.262) |  |  |  |  |
| gdppc |  | .0001285***<br>(0) |  | .0001242***<br>(0) |  | .0001242 ***<br>(0) |  | .0000992***<br>(0) |  | .0000903**<br>(0) |  | .0000968**<br>(0) |  | .0001085***<br>(0) |
| covid19_cases |  | 0<br>(0) |  | 0<br>(0) |  | 0<br>(0) |  | 0<br>(0) |  | 0<br>(0) |  | 0<br>(0) |  | 0<br>(0) |
| PRE_YEAR_C6M |  |  | -.405<br>(.635) | .477<br>(.607) |  |  |  |  |  |  |  |  | -1.034<br>(.725) | -.078<br>(.794) |
| C6M |  |  | 4.573<br>(4.302) | .341<br>(4.37) |  |  |  |  |  |  |  |  | 3.624<br>(4.344) | -3.302<br>(4.502) |
| POST_YEAR_C6M |  |  | .314<br>(.33) | -.093<br>(.334) |  |  |  |  |  |  |  |  | .619<br>(.385) | .179<br>(.447) |
| PRE_YEAR_C2M |  |  |  |  | -1.212<br>(.863) | -.972<br>(1.196) |  |  |  |  | -1.34<br>(.892) | -.28<br>(1.318) |  |  |
| C2M |  |  |  |  | 7.629<br>(5.249) | 1.625<br>(7.17) |  |  |  |  | 7.14<br>(5.325) | -1.544<br>(7.183) |  |  |
| POST_YEAR_C2M |  |  |  |  | .893**<br>(.416) | .91*<br>(.55) |  |  |  |  | .884*<br>(.464) | .6<br>(.711) |  |  |
| PRE_YEAR_CATCHUP |  |  |  |  |  |  | 1.047<br>(.708) | 1.59**<br>(.708) | .442<br>(1.097) | 1.642<br>(1.336) | .568<br>(1.508) | 2.097<br>(1.631) | -.585<br>(.811) | .07<br>(.874) |
| CATCHUP |  |  |  |  |  |  | -13.97***<br>(4.322) | -14.792***<br>(4.597) | -15.018***<br>(4.381) | -15.019***<br>(4.635) | -13.741***<br>(4.401) | -15.044***<br>(4.603) | -13.698***<br>(4.463) | -15.298***<br>(4.827) |
| POST_YEAR_CATCHUP |  |  |  |  |  |  | -.358<br>(.322) | -.533<br>(.331) | .646<br>(.417) | .408<br>(1.005) | -.323<br>(.665) | -.791<br>(.743) | .419<br>(.483) | .149<br>(.535) |
| PRE_YEAR_C1M_CA~P |  |  |  |  |  |  |  |  | .808<br>(1.137) | -.033<br>(1.357) |  |  |  |  |
| POST_YEAR_C1M_C~P |  |  |  |  |  |  |  |  | -1.18**<br>(.465) | -1.02<br>(1.005) |  |  |  |  |
| PRE_YEAR_C2M_CA~P |  |  |  |  |  |  |  |  |  |  | .581<br>(1.471) | -.712<br>(1.578) |  |  |
| POST_YEAR_C2M_C~P |  |  |  |  |  |  |  |  |  |  | -.009<br>(.664) | .435<br>(.729) |  |  |
| PRE_YEAR_C6M_CA~P |  |  |  |  |  |  |  |  |  |  |  |  | 2.418**<br>(1.006) | 2.445**<br>(1.024) |
| POST_YEAR_C6M_C~P |  |  |  |  |  |  |  |  |  |  |  |  | -1.136**<br>(.55) | -1.044*<br>(.589) |
| _cons | 74.571***<br>(5.966) | 79.107***<br>(10.124) | 77.524***<br>(3.612) | 78.87***<br>(3.829) | 74.815***<br>(4.816) | 77.723***<br>(6.753) | 84.406***<br>(2.172) | 83.875***<br>(2.528) | 76.449***<br>(6.274) | 85.648***<br>(10.907) | 78.822***<br>(5.138) | 85.321***<br>(7.108) | 81.805***<br>(4.148) | 86.387***<br>(4.589) |
| Observations | 1060 | 900 | 1060 | 900 | 1060 | 900 | 1060 | 900 | 1060 | 900 | 1060 | 900 | 1060 | 900 |
| Pseudo R² | .z | .z | .z | .z | .z | .z | .z | .z | .z | .z | .z | .z | .z | .z |

Standard errors are in parentheses \*\*\* p<.01, \*\* p<.05, \* p<.1

## Discussions

The substantial declines in PCV and other routine immunization coverage caused by the COVID-19 pandemic and its associated containment measures as well as their persistent lagging below pre-pandemic levels years after lockdown policy removal demonstrate that many countries lacked a level of health systems preparedness needed prevent routine health service disruptions. Key factors leading to this to decline in immunization coverage included pandemic lockdown policies, supply chain disruptions, shortages of healthcare workers, and vaccine hesitancy, which in turn led to more global outbreaks of vaccine-preventable diseases, such as measles and pneumococcal infections. Our study analyzes the impact of three pandemic lockdown policies (school closures, workplace closures, and stay-at-home orders) and the influence of immunization system preparedness as proxied by recent experience with pre-pandemic vaccine catch-up campaigns on PCV coverage levels and persistence of coverage declines in the post-pandemic period.

Consistent with prior literature, our findings demonstrate the positive impact vaccination catch-up campaigns including through mobile clinics, school-based and community-based campaigns, and mass vaccination programs in the pre-pandemic period (Kumar et al., 2025; Mancarella et al., 2022). We hypothesized that countries who successfully built capacity and rolled out these campaigns between 2018 and 2020 would have been more prepared to enact future catch-up campaigns to mitigate the impact of COVID-19 related disruptions on routine immunization. For example, Kumar et al. (2025) demonstrate that PCV vaccine catch-up campaigns, implemented by India’s Ministry of Health in 2021, increased the country’s post-pandemic national vaccination rate to greater than 80% by 2024. Mancarella et al. (2022) show that simultaneously implementing school-based and community-based vaccination campaigns and mobile clinics in Italy increased the national immunization coverage of Diphtheria-Tetanus-acellular Pertussis and Poliomyelitis vaccination (DTaPP4) from 63.5% to 73.1%. Furthermore, in Nigeria, a country with historically low coverage of pneumococcal conjugate vaccine (PCV), implementing post-pandemic mass campaigns targeted toward various age groups between 2021 and 2022 decreased nasopharyngeal pneumococcal carriage by approximately 16% among Nigerian children between age 1 and 9 years (Coldiron et al., 2025).

However, in contrast with some of these specific cases, our research shows that capacity developed in the pre-pandemic period through recent experience with vaccine campaigns and pandemic lockdown policies does not adequately protect or prepare countries from suffering large setbacks to routine immunization. Other systemic factors such as the resiliency of health systems and sufficient public health financing help ensure the timely delivery of routine immunizations during and after pandemics. Specifically, higher-income countries had more fiscal capacity in place to quickly roll out essential vaccination programs and adapt their vaccine campaign strategies to the challenges posed by the COVID-19 pandemic than lower-income countries, despite having similar capacity to implement catch-up campaigns. As a result, emergency financing for routine service catch-up should be considered as a part of future pandemic response strategy.

In conclusion, COVID-19 adversely impacted PCV vaccination coverage, and the longevity of this adverse impact was effect modified by the type of pandemic mitigation policy strategy employed by the country. Stay-at-home orders and school closures did not do anything to mitigate the effect of COVID-19 on PCV-coverage while countries enacting workplace closures only saw faster recovery of PCV coverage post-2020 toward pre-pandemic levels, than countries that did not enact workplace closures. As a result, policy impacting working adults, such as paid sick leave or targeted workplace closures in the face of a future pandemic or disaster, are likely to be the least disruptive types of pandemic response measures with respect to routine immunization services such as PCV vaccination.

Although recent catchup campaigns in the pre-COVID period were successful at increasing PCV coverage in the pre-period, the capacity built from enacting these campaigns neither mitigated the adverse impact of COVID-19 or COVID-19 control strategies on PCV coverage nor resulted in a faster recovery of PCV vaccination coverage toward pre-pandemic levels. This suggests that experience, preparedness to vaccinate, and capacity to deploy large-scale campaigns are unlikely to be sufficient to mitigate the impacts of future pandemic or disaster related disruptions to routine immunization. Health systems preparedness measures including supply chain security, adequate health workforce, and sufficient financial resources are likely needed in addition to campaign deployment capacity and training to mitigate the impacts that a future pandemic or disaster may have on routine immunizations such as COVID-19.

## Data Availability

All data produced in the present study are available upon reasonable request to the authors

## Appendix

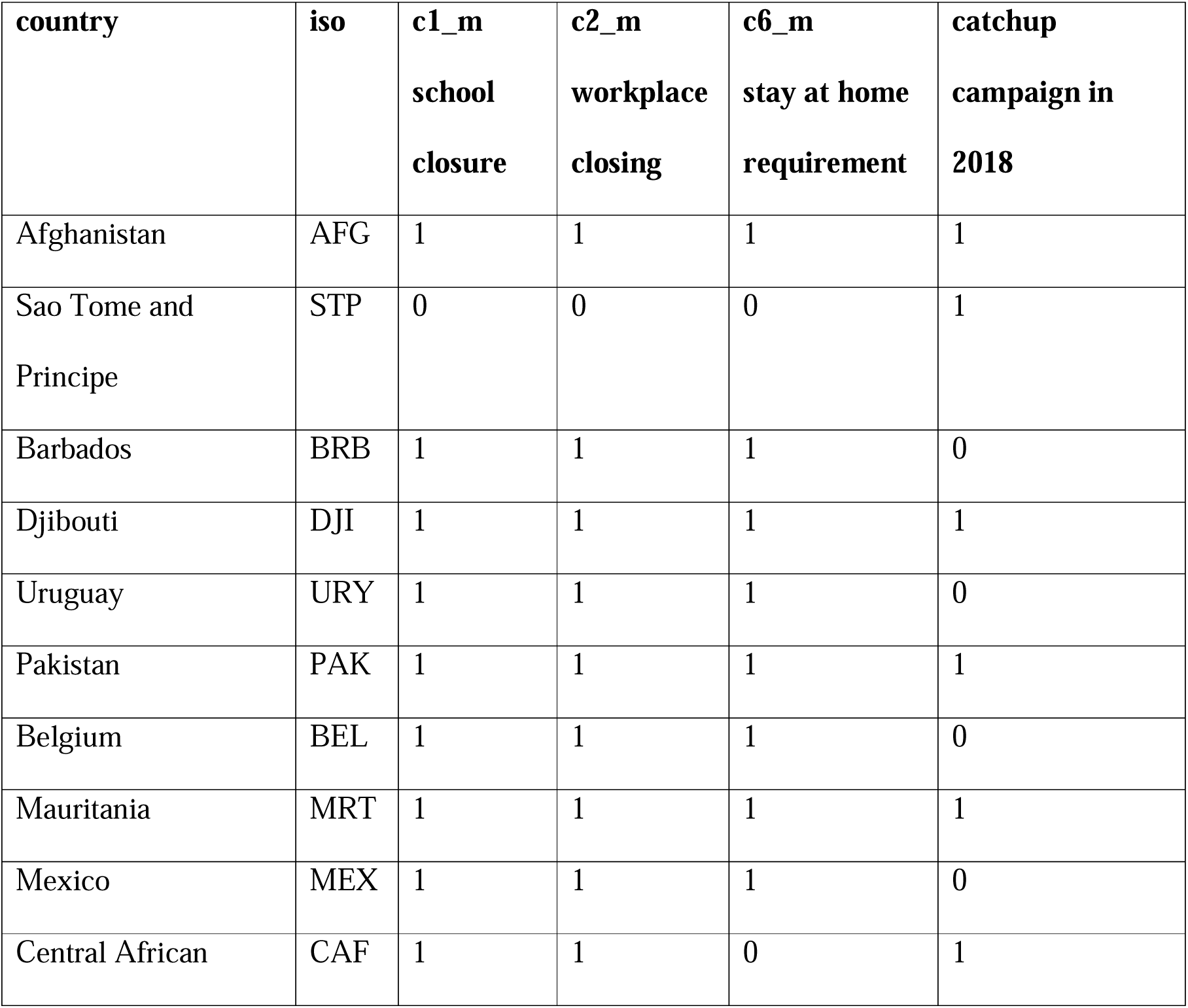
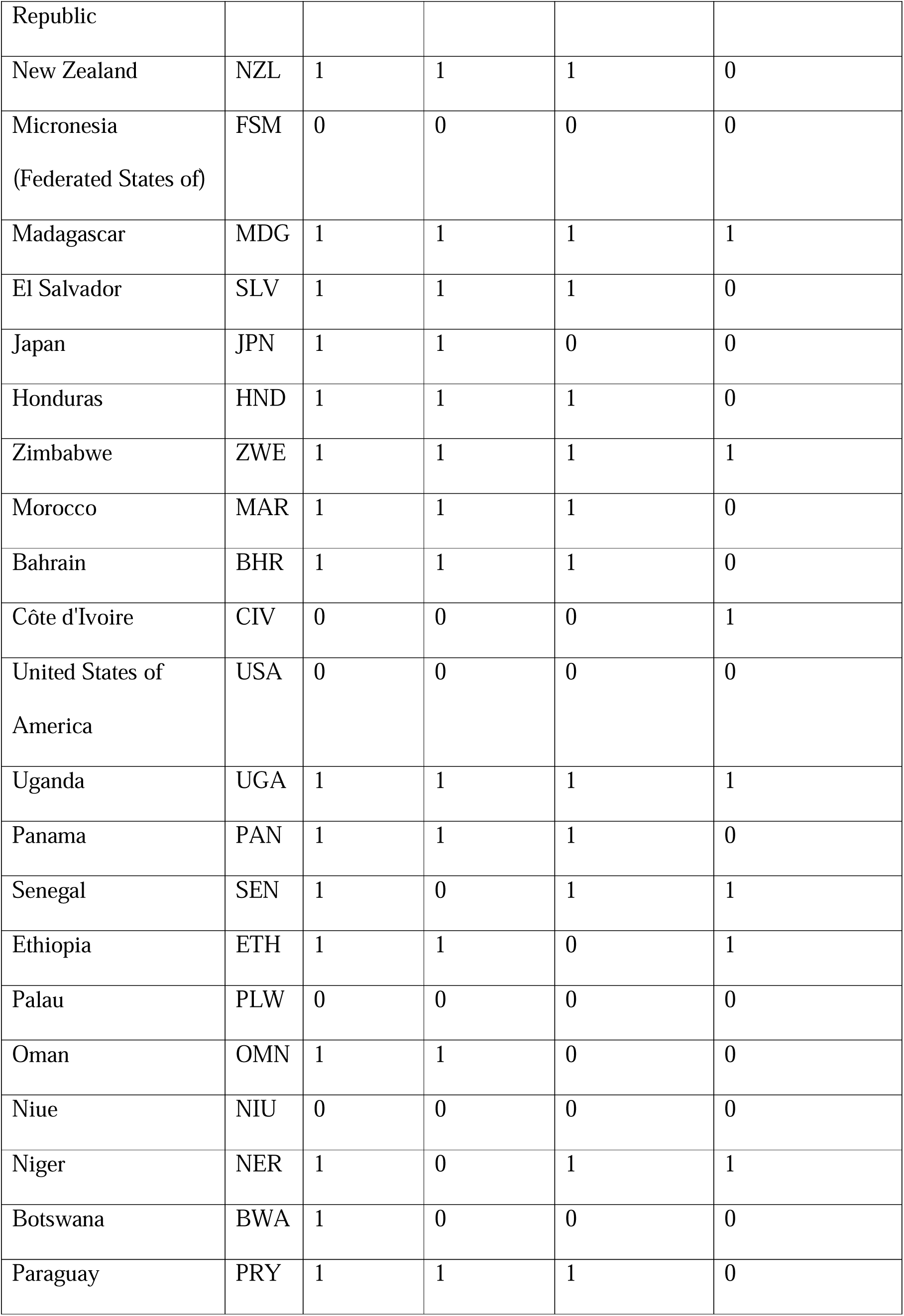
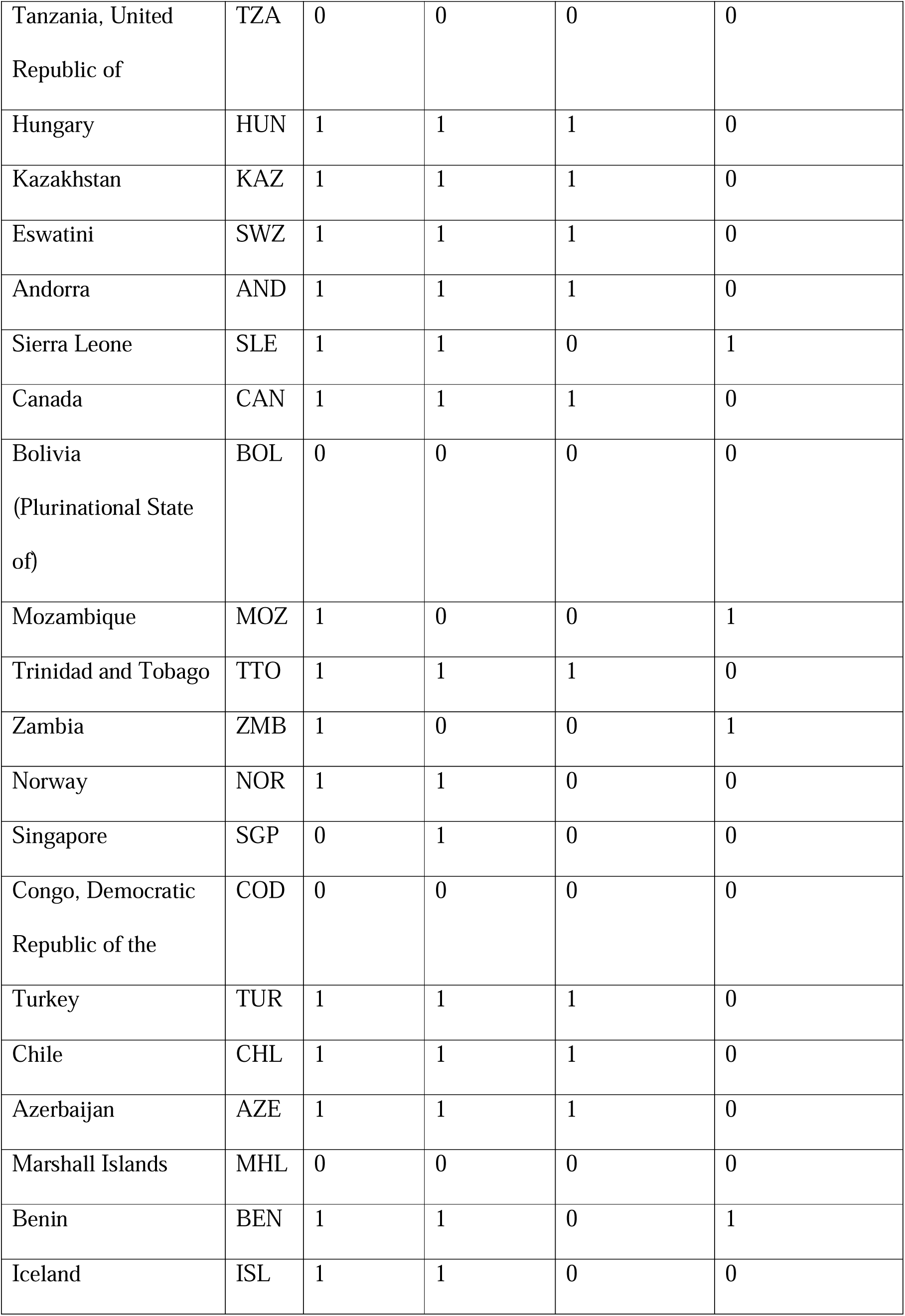
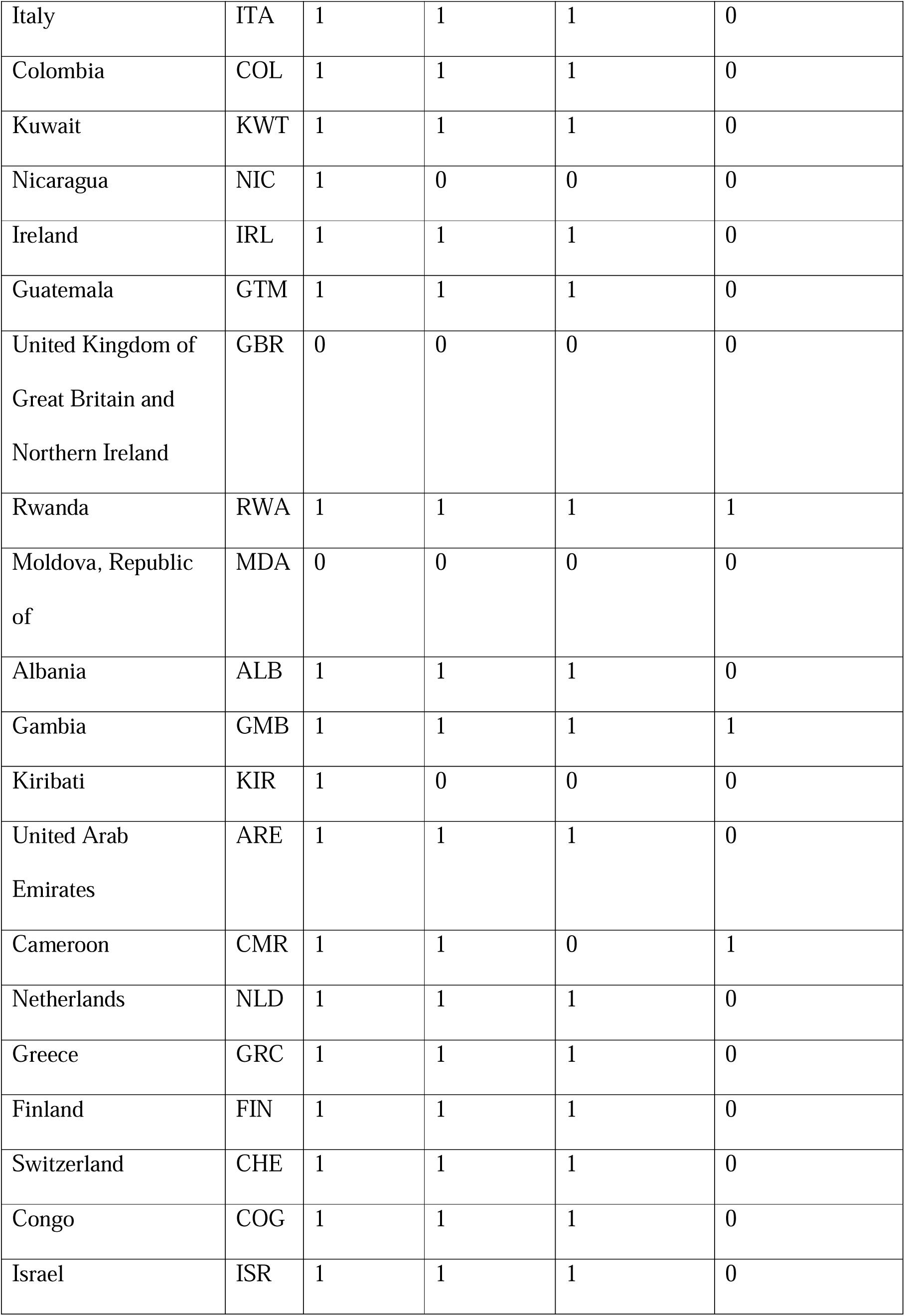
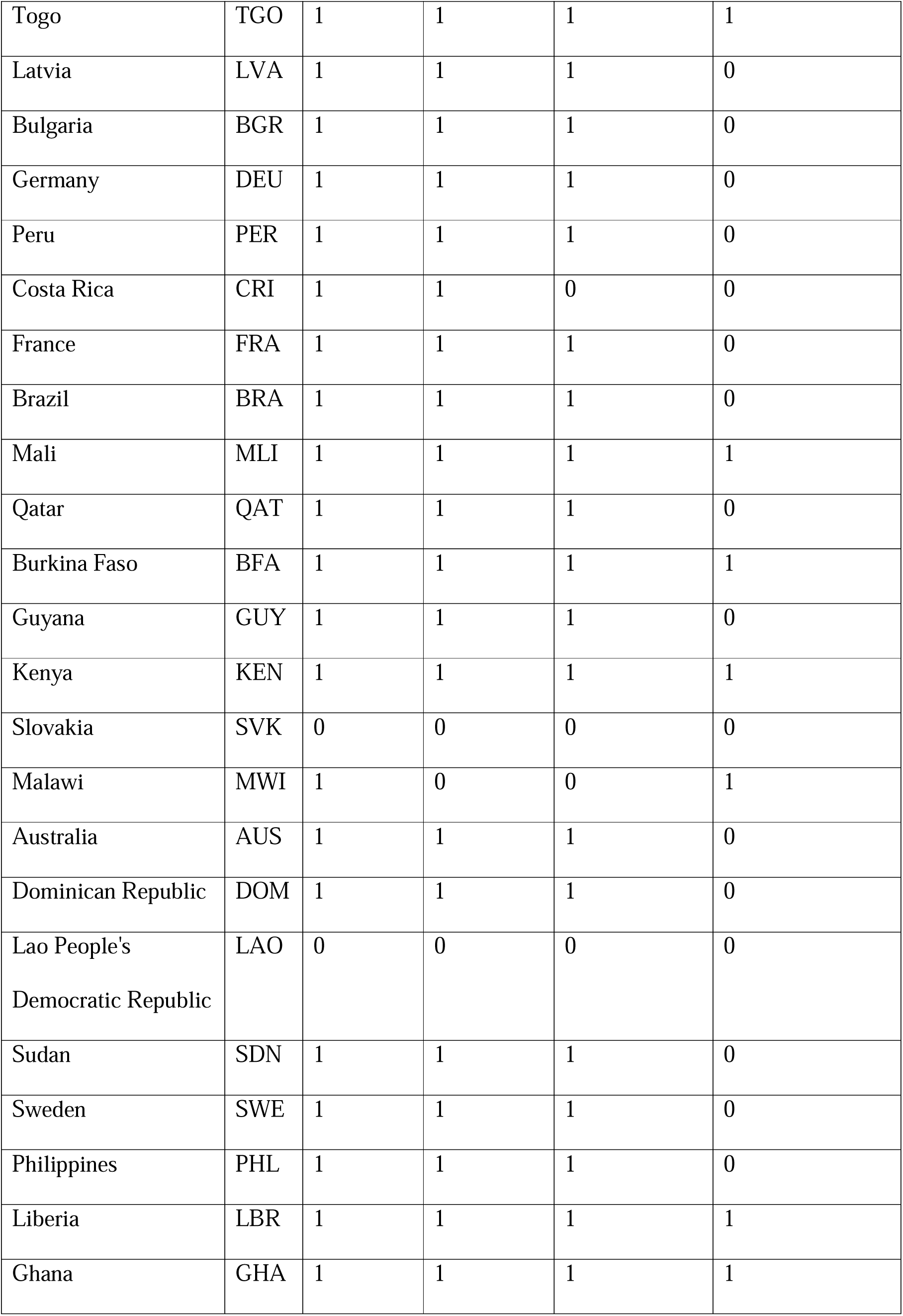
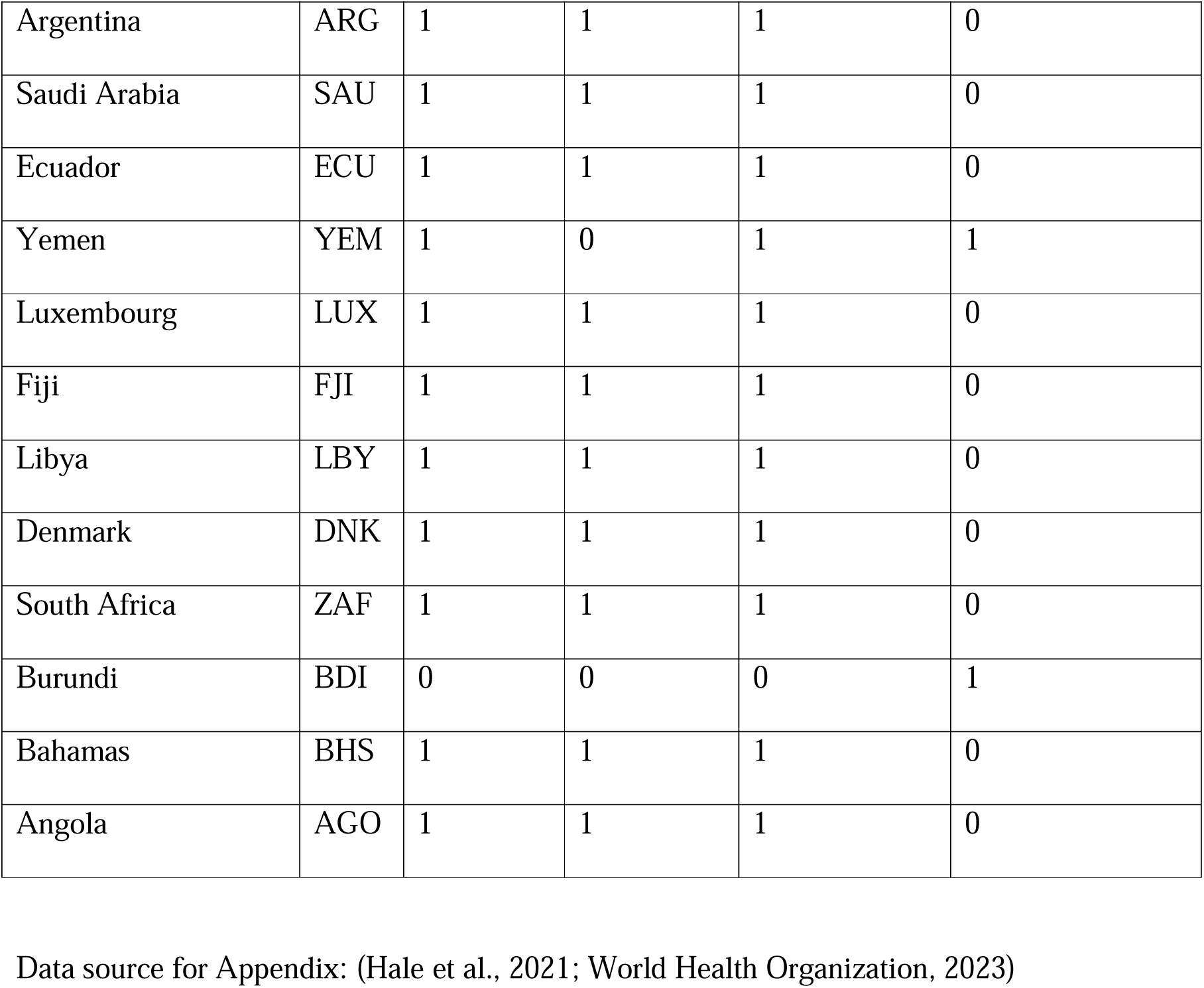

